# Emergence of a novel reassorted high pathogenicity avian influenza A(H5N2) virus associated with severe pneumonia in a young adult

**DOI:** 10.1101/2025.11.21.25340167

**Authors:** Joel Armando Vázquez-Pérez, Eduardo Becerril-Vargas, José Ernesto Ramírez-González, Mario Solís-Hernández, Charles Todd Davis, Pamela Garciadiego-Fossas, Marco Villanueva-Reza, Hansel Hugo Chávez-Morales, Enrique Mendoza-Ramírez, Christian Diego Olivares-Flores, América Citlali Vera-Jiménez, Uriel Rumbo-Nava, Cristóbal Guadarrama-Pérez, Elio Germán Recinos-Carrera, Joaquín Zúñiga, Irma López-Martínez, Lucía Hernández-Rivas, Gisela Barrera-Badillo, Nohemí Colin-Soto, Laura Flores-Cisneros, Guillermo Carbajal-Sandoval, Diana Vidal-Aguirre, Dayanira Sarith Arellano-Suarez, Rodrigo Aparicio-Antonio, Ramiro López-Elizalde, Carlos Javier Alcazar-Ramiro, Armando García-López, Han Di, Marie Kirby, Yunho Jang, Miguel Angel Lezana-Fernández, Carmen Margarita Hernández-Cárdenas

**Author notes:** Corresponding author: Carmen Margarita Hernandez-Cardenas Direccion General, Instituto Nacional de Enfermedades Respiratorias. Calzada de Tlalpan 4502, Belisario Domínguez Secc 16, Tlalpan, 14080, Ciudad de México.

## Abstract

**Background:** Infection of backyard and poultry with low pathogenicity avian influenza LPAI A(H5N2) viruses has occurred in Mexico since 1994, and the first human infection caused by this influenza virus was detected in 2024. Since its emergence in the Americas, frequent reassortments between high pathogenicity avian influenza HPAI A(H5N1) and LPAI viruses has occurred. In September 2025, the Instituto Nacional de Enfermedades Respiratorias of Mexico City identified an unsubtypeable influenza A virus infection in a young adult patient later determined to be a reassortant HPAI (H5N2) virus with a clade 2.3.4.4b HA.

**Methods:** We analyzed clinical and epidemiologic data from this patient. Respiratory samples were tested for influenza RT-qPCR assays. Genomic sequence and phylogenetics analyses were performed to provisionally assign a new genotype to the novel HPAI A(H5N2) reassortant virus.

**Results:** The patient presented with fever and tachypnea, later developed hemoptysis and thoracic pain, with oxygen saturation decreasing to 70%. CT scan showed bilateral ground-glass opacities consistent with diffuse alveolar hemorrhage and zones consistent with consolidation. Clinical improvement was observed and the patient was discharged.

Through viral complete genome analysis, we identified an HPAI A(H5N2) virus with genes from both clade 2.3.4.4b A(H5N1) viruses similar to those detected in North America during 2022-2023 and genes from the LPAI A(H5N2) viruses detected in Mexico during 2024.

**Conclusions:** This is the first ever laboratory-confirmed human infection caused by an HPAI A(H5N2) virus infection, suggesting a new genotype provisionally classified as B3.14. The relationship of the virus with the severity of illness remains unknown.

## Introduction

The high pathogenic avian influenza (HPAI) A H5N1 clade 2.3.4.4b virus was detected in North America in late 2021 (1). Since its emergence in the Americas, it has demonstrated the capacity to infected a wide spectrum of avian and mammalian species (2). Sporadic infections in humans have been detected after close contact with infected animals and a broad spectrum of disease from mild respiratory illness to sever and fatal outcome has been described (3). Since the introduction of clade 2.3.4.4b A(H5N1) virus into the Americas frequent reassortment and acquisition of low pathogenicity avian influenza (LPAI) viruses genes, including neuraminidase (NA) segments other than N1, have occurred resulting in numerous genotypes (4). The presence of other avian influenza viruses (IAV) in wild birds and poultry throughout the Americas continues to result in newly described HPAI A(H5) genotypes (5)(6). In Mexico, LPAI A(H5N2) viruses have been detected since 1994 and remain enzootic in poultry (7). In 2024, two LPAI A(H5N2) outbreaks were reported in backyard poultry in the State of Mexico in the metropolitan area near Mexico City (8). Moreover, in April 2024, a case of unsubtypeable influenza A virus was identified in a immunocompromised patient with renal failure (9). Sequencing of M, NS, NA, NP, and HA complete gene segments, identified an LPAI A(H5N2) virus with 99% identity to the A(H5N2) poultry outbreak viruses detected in the State of Mexico during 2024 (9). This case was the first ever reported human infection caused by a LPAI A(H5N2) virus in the world. In March 2025, Mexico reported the countrýs first case of HPAI (H5N1) in Durango State. This A(H5N1) genotype D1.1 virus infection resulted in a fatal outcome (10). In the present work, we describe the third case of AIV A(H5) infection in Mexico; a case of respiratory illness caused by a reassorted clade 2.3.4.4b HPAI A(H5N2) virus in a patient who was admitted to the Instituto Nacional de Enfermedades Respiratorias (INER) in Mexico City in late September 2025. We describe the atypical presentation of respiratory disease and the molecular characterization and classification of a new clade 2.3.4.4b HPAI A(H5N2) virus genotype.

## Methods

### Laboratory diagnostic and respiratory samples

An oropharyngeal swab and bronchoalveolar lavages (BAL) were obtained from patients. The initial diagnostic was performed using oropharyngeal swabs analyzed with the BioFire® Respiratory Panel 2.1. Further subtyping using the U.S. Centers for Disease Control and Prevention (CDC) seasonal influenza subtyping assay and finally, an RT-qPCR assay for avian influenza H5 (LightMix® Modular Influenza A H5, Roche) was performed on a BAL sample. Demographic characteristics, severity scores, comorbidities, antibiotic/medication exposure, and clinical outcome were collected for the patient.

### Influenza whole genome sequencing

The eight viral genome segments were amplified simultaneously from the clinical sample (BAL) using the universal primers MBT-Uni 12 and MBT-Uni 13, and libraries for the eight segments were sequenced on a MiSeq platform (Illumina, San Diego, CA, USA). The DRAGEN COVIDSeq Targeted Microbial Pipeline on BaseSpace Sequence Hub was used for analysis, mapping, and consensus sequence assembly. MEGAHIT was used to perform de novo assembly on the scrubbed reads. CD-HIT-EST was used to cluster similar contigs to reduce redundancy. The resulting contigs were mapped to a set of reference genomes using minimap2. Blast online (https://blast.ncbi.nlm.nih.gov/Blast.cgi) was used to assess the identities of viral consensus sequences.

Sequences of the eight gene genome segments of the HPAI A(H5N2) sample, INER_INF1727_25 were deposited in GISAID (EPI_ISL_20215425).

### Phylogenetic analysis

To perform phylogenetic analysis, we analyzed 225 complete genome sequences of avian influenza A(H5N2) viruses available on the GISAID platform from different states in Mexico (1994-2024) and North America as previously reported (9) to analyze the NA, NP and PB1 segments, which did not Blast to genes from HPAI A(H5N1) viruses. Additionally, 1830 complete genome sequences of avian influenza A(H5N1) viruses available on the GISAID platform from North America (2022-2025) were used to analyze M, NS, HA, PA and PB2 segments. As previously reported (9), sequence alignments were created using MAFFT V7 (11) and edited with MEGA 10.0 (12). A maximum likelihood tree was constructed for the whole genome sequence using MEGA 10.0. The General Time-Reversible model was selected with five-parameter gamma-distributed rates and 1000 bootstrap replicates. Edition of the trees was made using FigTree (13). For the phylogenetic analyses, we used two sequences obtained from a recent HPAI A(H5) virus outbreak during September 2025 in non-poultry birds in an urban area in Nezahualcóyotl, State of México (14).

To classify the genotype of the reassortant A(H5N2) virus from the case, we used GenoFLU software (15), setting the threshold to determine the genotype at >97% identity.

## Results

### Clinical case

A young adult female patient presented to the INER in Mexico City on late September 2025, with an 8-day history of sore throat, fever, cough, dyspnea, and hemoptysis. She reported no history of influenza vaccination. Her medical history was notable for obesity (body-mass index, 31.9) and close contact with birds, including poultry and pigeons; she had no history of travel or other relevant exposures.

The patient’s symptoms began with odynophagia and fever (temperature, 39°C). She initially sought care from a private practitioner and was prescribed antibiotics, but her condition did not improve. Lately, she had developed hemoptysis and thoracic pain, prompting admission. On presentation, physical examination revealed an oxygen saturation of 75% while breathing ambient air. Arterial blood gas analysis showed severe hypoxemia, with a PaO₂:FiO₂ ratio of 88 and an elevated alveolar–arterial oxygen gradient. Supplemental oxygen via nasal cannula was initiated, resulting in clinical stabilization.

Laboratory evaluation demonstrated a normal white-cell count, an elevated lactate dehydrogenase level (1241 U/L), and an elevated C-reactive protein level (38.2 mg/L). Chest computed tomography (CT) revealed bilateral ground-glass opacities with subpleural and axial distribution, interlobular septal thickening, a crazy-paving pattern, alveolar filling, and air bronchograms, predominantly affecting the right lung (Figures 1).

**Figure 1.**
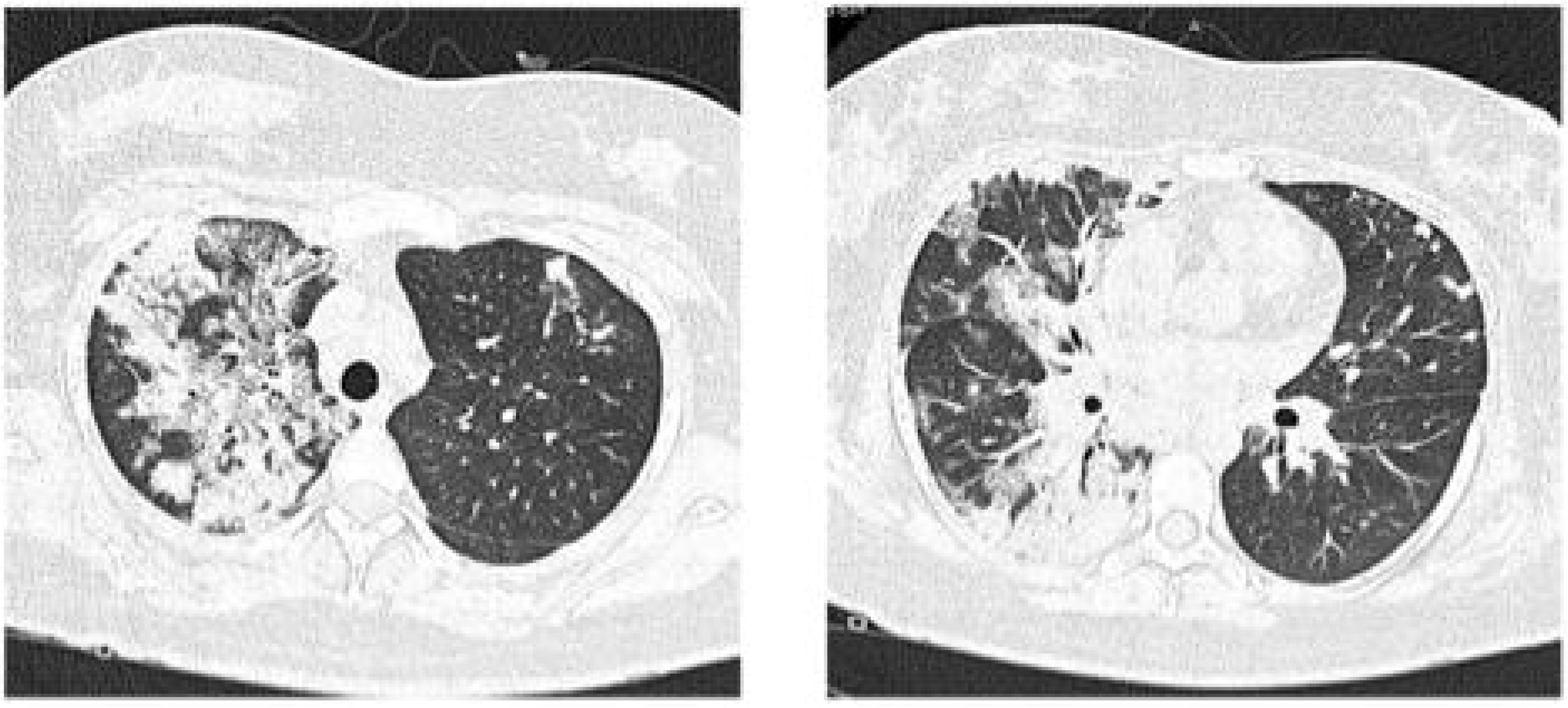
CT scan upon admission—Left: The right upper lobe demonstrates ground-glass opacities and interlobular septal thickening with a crazy-paving pattern, predominantly with subpleural and axial distribution. Multiple solid nodules are observed in the anterior segment of the left upper lobe. Right: Right lung: Consolidation with air bronchograms, ground-glass opacities, and a crazy-paving pattern involving the middle and lower lobes, predominantly with subpleural and axial distribution. Left lung: Several solid nodules with subpleural distribution located in the lingula.

Initial diagnostic testing of a oropharyngeal swab sample indicated positivity for influenza A virus. The respiratory pathogen panel of a bronchioalveolar lavage (BAL) sample was positive for influenza A virus, with no subtype identified. Further testing using the CDC seasonal influenza typing/subtyping protocol showed amplification for influenza A (Ct, 31.06), but the subtype remained undetermined. A RT-qPCR assay for avian influenza H5 on the BAL sample was positive, with a Ct of 32.7.

The patient was admitted with a diagnosis of severe atypical pneumonia, with a MuLBSTA score of 9 points, a PSI/PORT score of 58 points (Class II), and two minor criteria meeting IDSA/ATS severity criteria. Contact and droplet precautions were implemented pending further evaluation. As part of the differential diagnosis, autoimmune causes and immunosuppression were sought and ruled out.

Bronchoscopy with bronchoalveolar lavage (BAL) was performed and a hemorrhagic sample was obtained. Histopathologic examination of the biopsy specimen (Figure 2) revealed pneumocyte hyperplasia, microscopic foci of recent intra-alveolar hemorrhage, and lymphocytic infiltrate, with no evidence of vasculitis. The patient received supportive care, including supplemental oxygen via nasal cannula and treatment with oseltamivir at day 2 after admission. Her condition improved steadily, and she was discharged without complications. As part of the case investigation, contact tracing was implemented, with the health department conducting local visits to search for, birds including poultry, and other animals that could be infected with avian influenza A(H5), virus as well as family members and neighbors. Forty-one contacts were identified and samples were taken from the identified contacts and they were given oseltamivir prophylaxis, no respiratory symptoms were detected. No identified human contacts were found positive for influenza A(H5) (16). A dog was identified as a pet at the case’s residence, and several animals were found in the courtyard, including a poultry bird and two pigeons. Samples collected from the identified animals were tested positive for influenza A(H5) (16).

**Figure 2.**
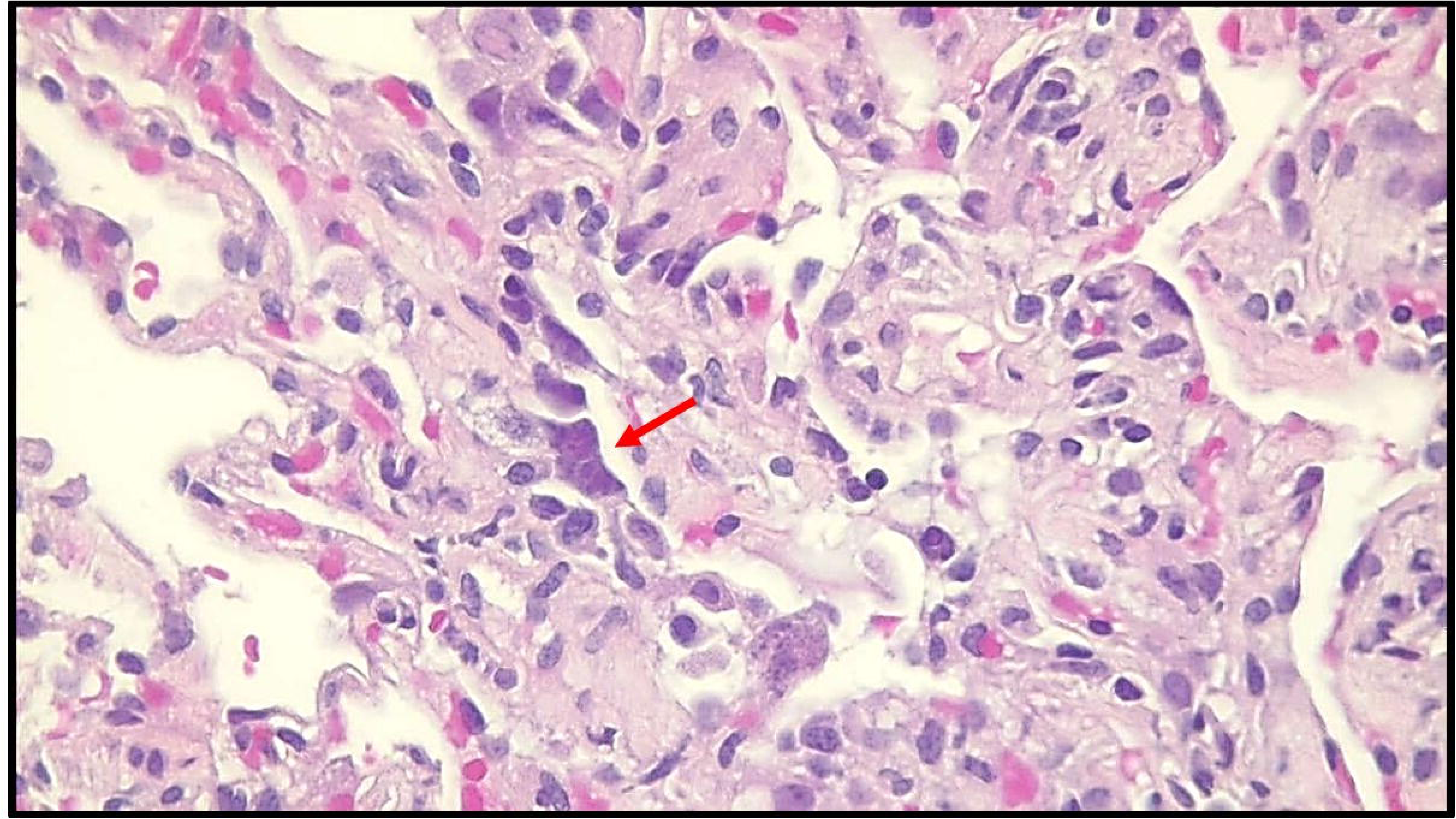
Microscopic photograph at 40x. In hematoxylin and eosin staining, red arrows indicate hyperplasia of type II pneumocytes, characterized by nuclear basophilia and a prominent nucleolus, which is secondary to cytopathic damage.

### Molecular Characterization and Phylogenetic Analyses of the A(H5) virus

Using a de novo assembly pipeline of 88,910 unique influenza-specific reads, we obtained each of the eight codon complete gene segments. The consensus sequence of each gene segment was assessed by BLAST analyses demonstrating 98% nucleotide sequence identity of the PB1, NP and NA genes with the LPAI A(H5N2) virus, A/State of Mexico/INER-INF645/2024, detected in 2024 (9). In contrast, PB2, PA, HA, M and NS genes showed 98% identities with sequences of HPAI A(H5N1) viruses detected in North-America during 2022-2023. The closest Blast hit compared to these genes were PB2: A/great-tailed grackle/Kansas/W22-1223B/2022 (A/H5N1), PA: A/blue-winged teal/Minnesota/AV22-675/2022 (A/H5N1), HA: A/Skunk/AB/FAV-0897-02/2022 (A/H5N1), M: A/blue-winged teal/Texas/UGAI22-3190/2022 (A/H5N1), NS: A/great-tailed grackle/Kansas/W22-1223B/2022 (A/H5N1). Each of the genes from the closest Blast hits were from A(H5N1) viruses classified as the B3.2 genotype (15).

Phylogenetic analysis showed that the sequences of segments NA (Figure 4), NP, and PB1 genes (supplemental material) clustered with the sequences of LPAI A(H5N2) from 2022, 2023, and 2024. Two sequences from the most recent outbreak from 2025 in the State of Mexico were included. These viruses were related to the high disease burden and mortality events in non-poultry birds in Nezahualcoyotl (14), a locality in the State of Mexico near the residence of the patient. The A(H5N2) human and avian NA sequences formed a robust distinct monophyletic group and tended to clustered with more recent sequences from Mexico from 2024 suggesting an LPAI A(H5N2) ancestor of the NA, NP and PB1 genes of the virus that caused the human case.

On the other hand, phylogenetic analysis showed the sequences of segments PB2, PA, HA, M and NS clustered with the genes of clade 2.3.4.4b A(H5N1) viruses detected in North America during 2022 and 2023. Phylogenetic analysis of the HA gene showed that the human case formed a robust distinct monophyletic group with other HPAI A(H5N2) backyard viruses from the State of Mexico detected during a recent outbreak from 2025 (Figure 3). This monophyletic group, which formed a separate branch in the phylogenetic tree (Figure 3) clustered within a larger group of B3 genotype A(H5Nx) viruses that have circulated widely in wild birds in the Americas and that have been detected in avian and mammalian hosts since 2022 (Figure 3). Compared to A/Astrakhan/3212/2020(H5N8) as reference, there were several mutations in the HA signal peptide: K3T, V5I, V14G; HA1: M135V, P140S, A160T, T192I, P239Q and HA2:K503R (H3 mature HA numbering). Moreover, there were several non-synonymous mutations differences between the human and avian sequences: P127S, T131A, R173Q, P239Q, E489A in HA, N207D, S216P in NS1, I211V in NA, A152T, N510D, K521T, E627K, T676A in PB2, L598P in PB1 and A231T, D272G, R330I, E352G, S409G in PA. Phylogenetic analysis of PB2, PA, M and NS genes showed similar clustering patterns as the HA gene (supplemental material).

**Figure 3.**
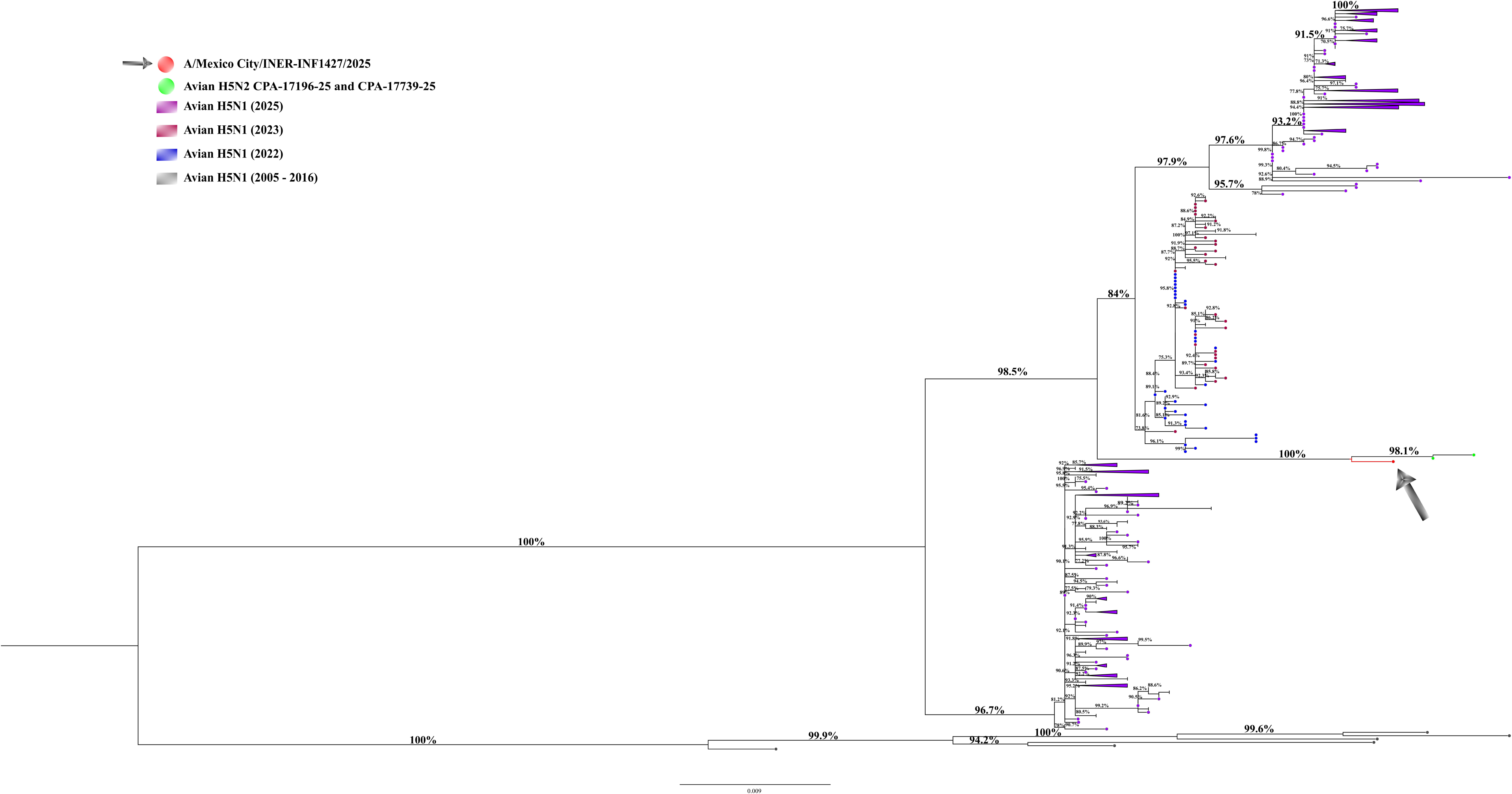
Maximum likelihood (ML) phylogenetic trees for HA influenza H5N1 genetic segment. ML trees from 1830 avian influenza H5N1 viruses (HPAIVs) from America registered in GISAID were produced with 1000 bootstrap replicates. The 2025 human sequence from Mexico is included (red circle and black arrow), along with sequences from Nezahualcoyotl (green circle). Avian sequences from different years are indicated with colored rectangles. Bootstrap values higher than 70% are indicated. The scale bar indicates the nucleotide substitutions per site.

**Figure 4.**
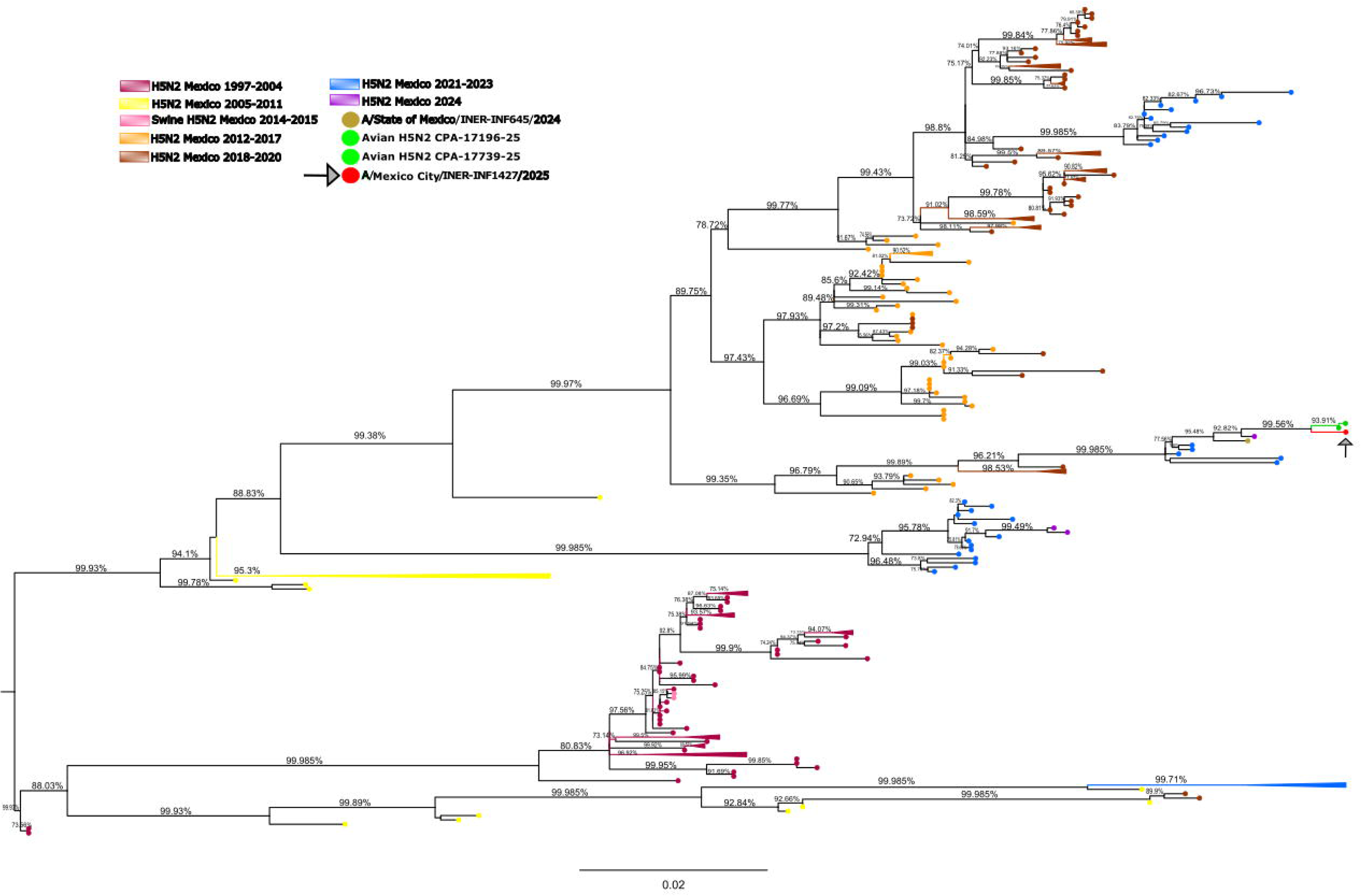
Maximum likelihood (ML) phylogenetic trees for NA influenza H5N2 genetic segments. ML trees from 225 avian influenza H5N2 viruses (LPAIVs) from America registered in GISAID were produced with 1000 bootstrap replicates. The 2025 human sequence from Mexico is included (red circle and black arrow), along with sequences from Nezahualcoyotl (green circle). Avian sequences from different years are indicated with colored circles. Bootstrap values higher than 70% are indicated. The scale bar indicates the nucleotide substitutions per site.

The multi-basic amino acids at the cleavage site of the HA showed five basic amino acids: PLREKRRKR/G, consistent with other HPAI A(H5Nx) viruses detected in the Americas. Mutations which confer resistance to oseltamivir, zanamivir, and peramivir in A(H5) viruses or HA molecular markers associated with increased binding to human-type receptors such as E190N, Q226L, and G128S (17) were also not present in this virus. However, we observed mutations at HA T192I and T199I in the receptor binding domain (RBD) which have been detected in some other A(H5) virus infected humans (18). The PB2 E627K mutation (40% K allele frequency) and K526R, both related with enhance polymerase activity and adaptation to mammalian cells, were also detected (19).

The genotype classification based on the reassortment of the 8 gene segments of the virus with respect to Euro-Asiatic (ea) A(H5Nx) ancestors was assessed using the GenoFlu tool (15). Using a 97% identity cutoff, GenoFlu classified the PB2, PA, HA, M and NS segments as belonging to the am2.1, ea1, ea1, ea1 and am1.1 lineages, respectively (Table 1). Each of these gene lineages were also detected in B3.2 genotype viruses from 2022-2023 and corresponded to the highest percent similarity gene segments of B3.2 viruses that were identified by Blast analysis. However, due to reassortment with genes derived from LPAI A(H5N2) subtype viruses, the NA, NP and PB1 segments do not belong to A/goose/Guangdong/1/96-lineage viruses resulting in a previously undescribed clade 2.3.4.4b HPAI A(H5N2) virus genotype. Using the GenoFlu genotyping criteria, HPAI A(H5) viruses originating from the A1 lineage (ea1) that have reassorted with North American LPAI viruses are classified as genotype B, and viruses originating from B3 viruses reassorted with LPAI viruses are further classified as B3.X (where X = the next number in the sequential numbering). According to these criteria, the data presented indicate a provisional classification of the virus as B3.14.

**Table 1.**
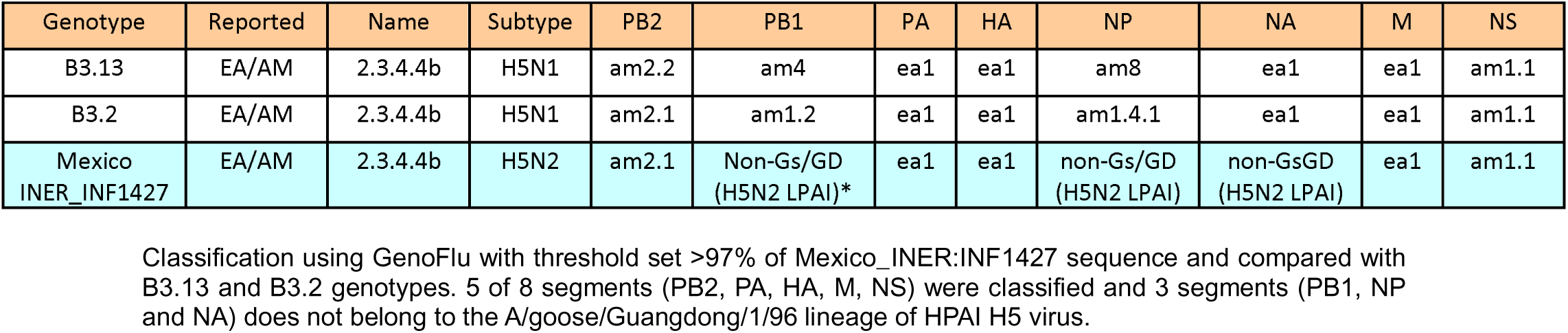
Genotyping of H5Nx human virus using GenoFlu.

## Discussion

This case illustrates a rare disease manifestation of an HPAI A(H5N2) virus infection in a young woman with close contact to birds, who presented with severe pneumonia complicated by hemoptysis. Previous reports have documented pulmonary hemorrhage as a potential complication of influenza pneumonia (20). Occasionally, H5 virus-infected patients develop severe pneumonia and acute respiratory distress syndrome (ARDS) characterized by diffuse alveolar damage, as observed in this case with hemorrhagic features.

In other reported case series, patients with avian influenza are often young adults who have been occupationally exposed to poultry through handling or proximity. Notably, most of these individuals lack underlying health conditions and present with symptoms such as high-grade fever, cough, and chest pain. Laboratory findings in these cases are similar to those in our patient, including normal white blood cell counts and elevated lactate dehydrogenase (LDH) levels in some instances (21). High-grade inflammation, driven by elevated interleukin levels, is a predominant feature in affected patients (22).

Mortality rates following infection with HPAI A(H5) are significant, reaching approximately 40% in some series despite treatment, and even higher in those not receiving oseltamivir (23)(24). This high mortality underscores the need for heightened collective efforts in early diagnosis and effective treatment to mitigate the impact of avian influenza.

Although human cases are relatively rare and the virus primarily affects birds, wild and domestic mammals have also tested positive for HPAI A(H5) and experienced severe morbidity and mortality in addition to sustained circulation of the virus in U.S dairy cattle (2). This highlights the potential for cross-species transmission and the importance of ongoing monitoring.

Viral sequence evidence suggests that the human virus (A/Mexico_City/INER_INF1427_2025) and the HPAI A(H5N2) avian viruses detected during 2025 represent reassortment between an enzootic LPAI A(H5N2) virus ancestor from 2024 detected in Central Mexico and the A(H5N1) clade 2.3.4.4b genotype B3.2 viruses detected during 2022-2023. Lack of more recently detected B3.2 genotype viruses containing genes with higher nucleotide similarity to the virus from the human case likely reflects undersampling or less sequencing of viruses that may be present in wild birds or poultry in the region. The highest nucleotide similarity of the PB1, NP and NA genes (98%) of the human virus compared to LPAI A(H5N2) viruses from avian and human hosts in 2024, suggests co-circulation of HPAI A(H5N1) and LPAI A(H5N2) viruses in the region resulting in reassortment (Figure 5). We hypothesized that the high disease burden of HPAI A(H5) viruses in chickens in this geographical area in September 2025 could be the reason of the human infection, although direct contact between the patient in this study and poultry or other domestic animals could not be confirmed,. Human-to-human transmission seems implausible and follow-up investigation of close contacts of the case did not identify other H5-positive individuals.

**Figure 5.**
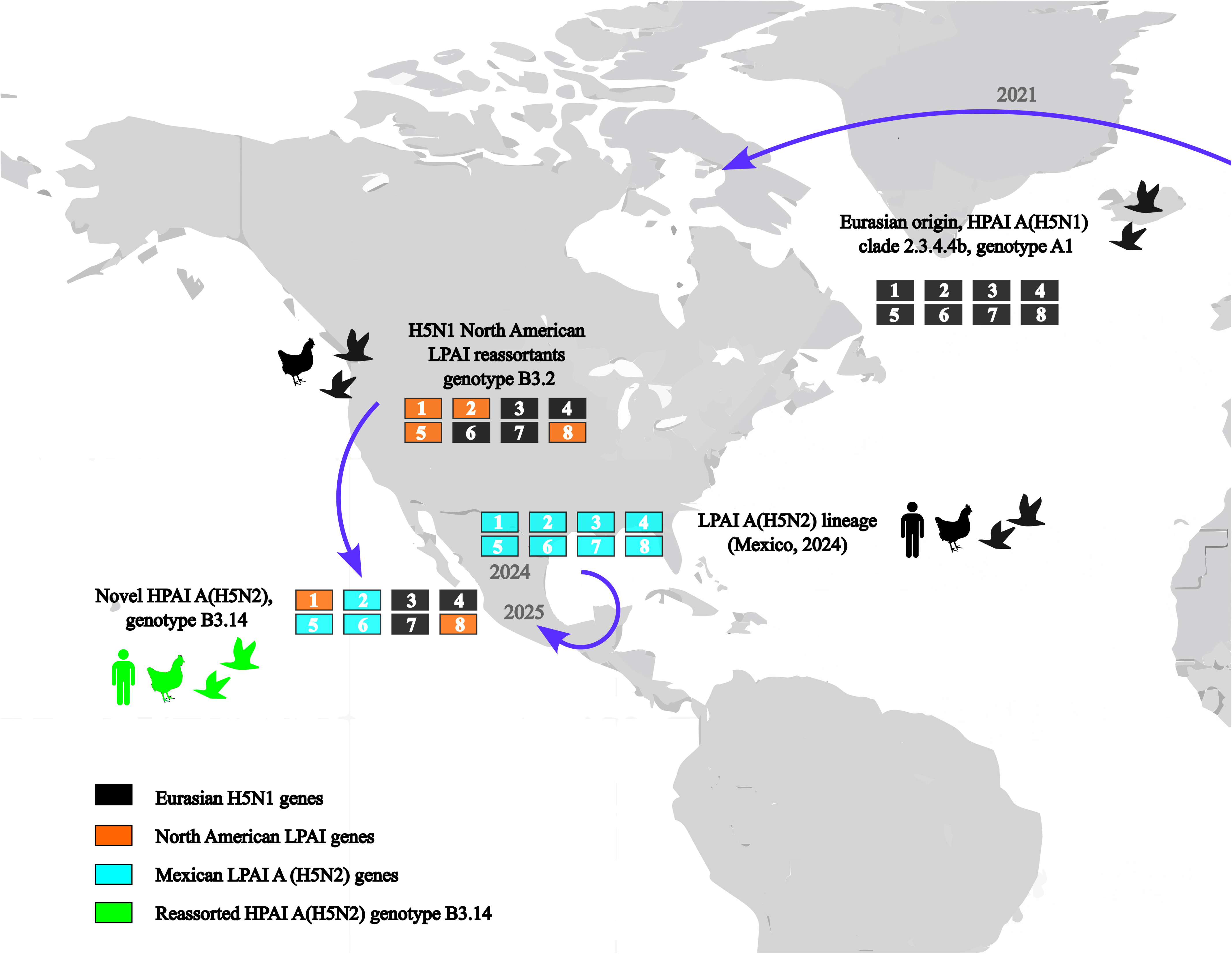
Reassortment events between the enzootic LPAI A(H5N2) virus ancestor from 2024 detected in Central Mexico and the A(H5N1) clade 2.3.4.4b genotype B3.2 viruses detected during 2022-2023. Each box represents one of the eight segments of the IAV genome. 1-PB2, 2-PB1, 3-PA, 4-HA, 5-NP, 6-NA, 7-MP, and 8-NS. Each box is shaded according to the AIV lineage of that segment, with black = Eurasian H5N1, orange = representing North American LPAIV, blue = representing North American LPAIV H5N2.

Further studies are required to determine the predicted pathogenicity and the transmissibility of the virus and its potential threat to human health. Although obesity was the sole comorbidity, the patient exhibited unusually extensive pulmonary damage, underscoring the need for further characterization of their pathogenic potential of this or related viruses. Since no cases of this reassorted A(H5N2) influenza virus in humans have been previously reported, we are unaware of the clinical outcomes that this HPAI virus subtype may have in humans. Given the virus’s propensity for rapid genetic reassortment, genomic surveillance is essential, particularly for emerging strains. Such surveillance forms a critical component of global preparedness and rapid response strategies, enabling countries to strengthen viral diagnostics, vaccine development and therapeutic strategies to prevent widespread outbreaks.

In summary, our findings support the emergence of a new clade 2.3.4.4b reassortant virus provisionally classified as genotype B3.14 and the first ever global human case of an HPAI A(H5N2) virus infection.

## Supporting information

Figure S1A and B. Maximum likelihood (ML) phylogenetic trees for PB2 (A) and PA (B) influenza H5N1 genetic segment. ML trees from 1830 avian influenza

Figure S1A and B. Maximum likelihood (ML) phylogenetic trees for PB2 (A) and PA (B) influenza H5N1 genetic segment. ML trees from 1830 avian influenza

Figure S2A and B. Maximum likelihood (ML) phylogenetic trees for M (A) and NS (B) influenza H5N1 genetic segment. ML trees from 1830 avian influenza H

Figure S2A and B. Maximum likelihood (ML) phylogenetic trees for M (A) and NS (B) influenza H5N1 genetic segment. ML trees from 1830 avian influenza H

Figure S3A and B. Maximum likelihood (ML) phylogenetic trees for PB1 (A) and NP (B) influenza H5N2 genetic segments. ML trees from 225 avian influenza

Figure S3A and B. Maximum likelihood (ML) phylogenetic trees for PB1 (A) and NP (B) influenza H5N2 genetic segments. ML trees from 225 avian influenza

---

Figure S1A and B. Maximum likelihood (ML) phylogenetic trees for PB2 (A) and PA (B) influenza H5N1 genetic segment. ML trees from 1830 avian influenza H5N1 viruses (HPAIVs) from America registered in GISAID were produced with 1000 bootstrap replicates. The 2025 human sequence from Mexico is included (red circle and black arrow), along with sequences from Nezahualcoyotl (green circle). Avian sequences from different years are indicated with colored circles. Bootstrap values higher than 70% are indicated. The scale bar indicates the nucleotide substitutions per site.

Figure S2A and B. Maximum likelihood (ML) phylogenetic trees for M (A) and NS (B) influenza H5N1 genetic segment. ML trees from 1830 avian influenza H5N1 viruses (HPAIVs) from America registered in GISAID were produced with 1000 bootstrap replicates. The 2025 human sequence from Mexico is included (red circle and black arrow), along with sequences from Nezahualcoyotl (green circle). Avian sequences from different years are indicated with colored circles. Bootstrap values higher than 70% are indicated. The scale bar indicates the nucleotide substitutions per site.

Figure S3A and B. Maximum likelihood (ML) phylogenetic trees for PB1 (A) and NP (B) influenza H5N2 genetic segments. ML trees from 225 avian influenza H5N2 viruses (LPAIVs) registered in GISAID were produced with 1000 bootstrap replicates. The 2025 human sequence from Mexico is included (red circle and black arrow), along with sequences from Nezahualcoyotl (green circle). Avian sequences from different years are indicated with colored circles. Bootstrap values higher than 70% are indicated. The scale bar indicates the nucleotide substitutions per site.

## Institutional Review Board Statement

This study was reviewed and approved by the Science, Biosecurity, and Bioethics Committee of the Instituto Nacional de Enfermedades Respiratorias (protocols numbers B22-23 and B37-25).

## Data Availability

The genomic information generated during the current study is available in GISAID database. Sequence of novel reassortment HPAIV H5N2 from Mexico were deposited in GISAID under accession number EPI_ISL_20215425. GISAID Identifier EPI_SET_251122hr. DOI: https://doi.org/10.55876/gis8.251122hr

## Acknowledgments

We thank Eduardo Márquez García from the Unidad de Biología Molecular, INER for technical assistance in Illumina sequencing. We also thank all physicians in the ICU for assistance with patient management, specially to Feliana Muñoz Reyes and Josue Cornejo Miranda.

## Informed Consent Statement

Informed consent was provided according to the Declaration of Helsinki. Written informed consent was obtained from the patient and/or from their relatives or authorized legal guardians prior to the publication of this paper.

## Author contributions

JAVP, EBV, JERG, ILM and CMHC conceived and designed the project. PGF, MVR, LHR, NCS, LFC, GCS, DVA, DSAS, RAA, RLE and MALF collected the clinical and epidemiological data. MSH, HHCM, EMR, CDOF, ACVJ, GBB, CJAR, AGL, HD, MK, YJ and JAVP performed the experimental laboratory procedures. ERT, CDOF, CTD and JAVP performed the bioinformatics and phylogenetic analyses. MVR, URN, CGP, EGRC and CMHC performed the interpretation of clinical data. EBV, JERG, CTD, MVR, JZ, CMHC and JAVP wrote the manuscript. JAVP and CMHC supervised the project and led the team. All authors have read and agreed to the published version of the manuscript.

