## Supplementary figures and images for "Emergence of a novel reassorted high pathogenicity avian influenza A(H5N2) virus associated with severe pneumonia in a young adult"

### Figure S1A and B. Maximum likelihood (ML) phylogenetic trees for PB2 (A) and PA (B) influenza H5N1 genetic segment. ML trees from 1830 avian influenza

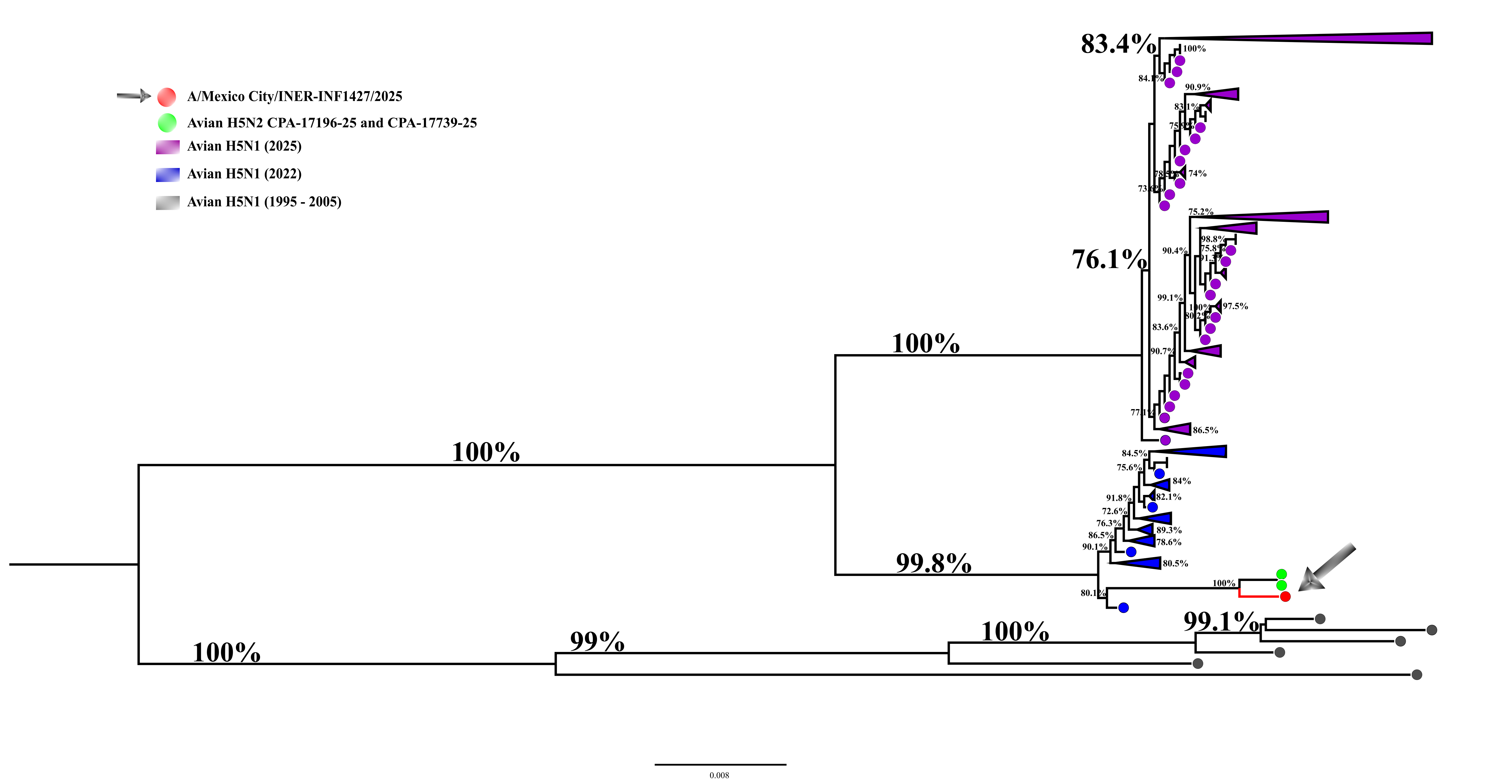

### Figure S1A and B. Maximum likelihood (ML) phylogenetic trees for PB2 (A) and PA (B) influenza H5N1 genetic segment. ML trees from 1830 avian influenza

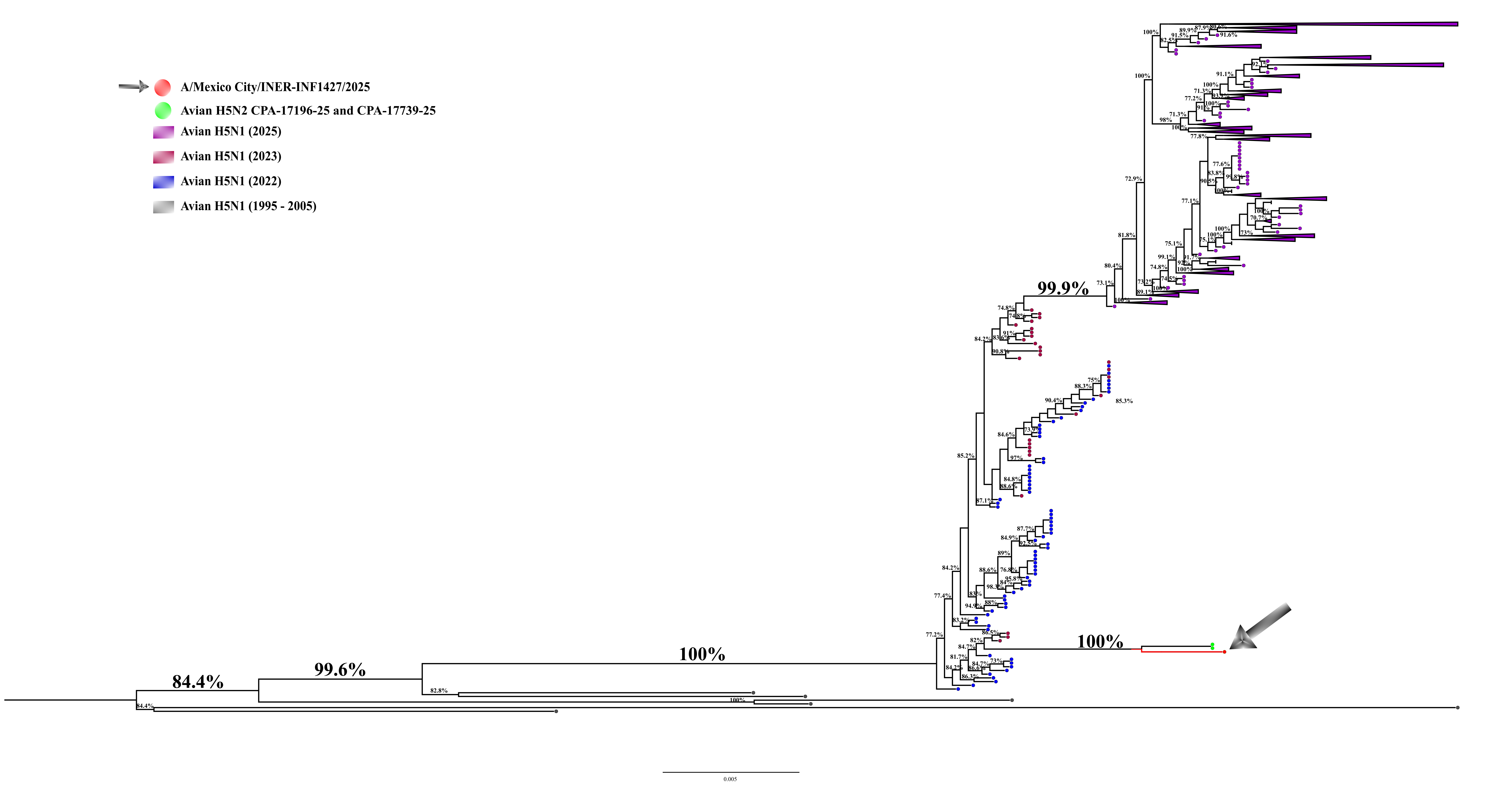

### Figure S2A and B. Maximum likelihood (ML) phylogenetic trees for M (A) and NS (B) influenza H5N1 genetic segment. ML trees from 1830 avian influenza H

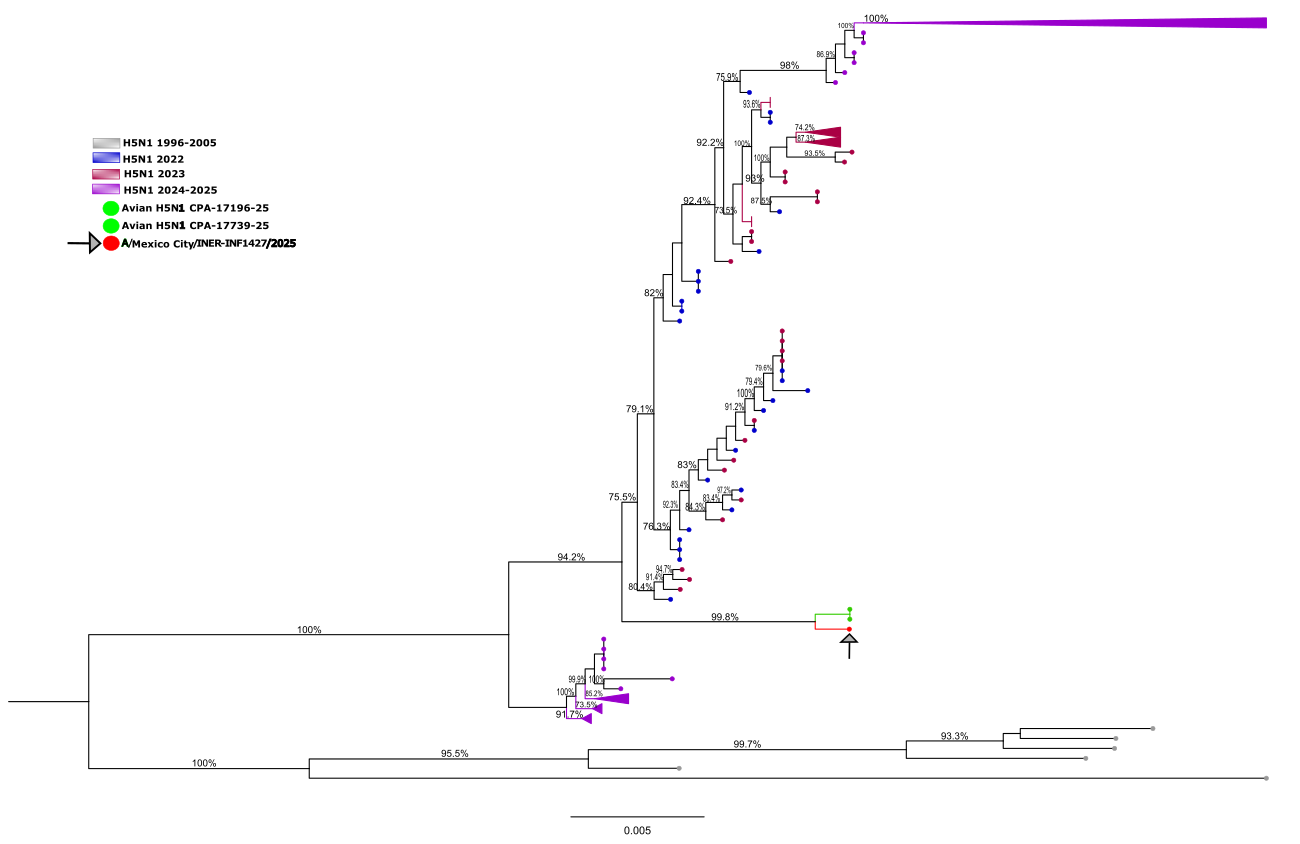

### Figure S2A and B. Maximum likelihood (ML) phylogenetic trees for M (A) and NS (B) influenza H5N1 genetic segment. ML trees from 1830 avian influenza H

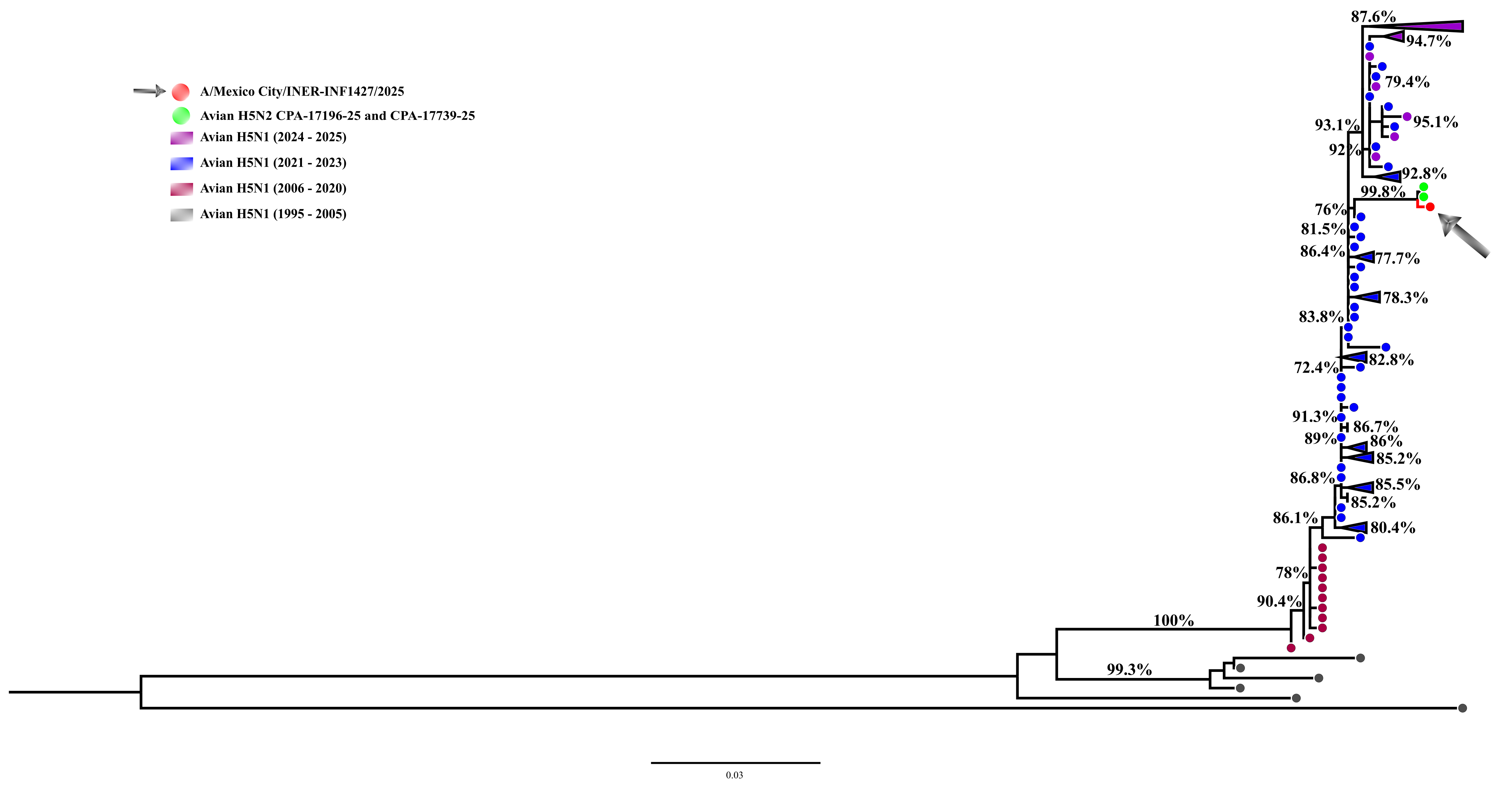

### Figure S3A and B. Maximum likelihood (ML) phylogenetic trees for PB1 (A) and NP (B) influenza H5N2 genetic segments. ML trees from 225 avian influenza

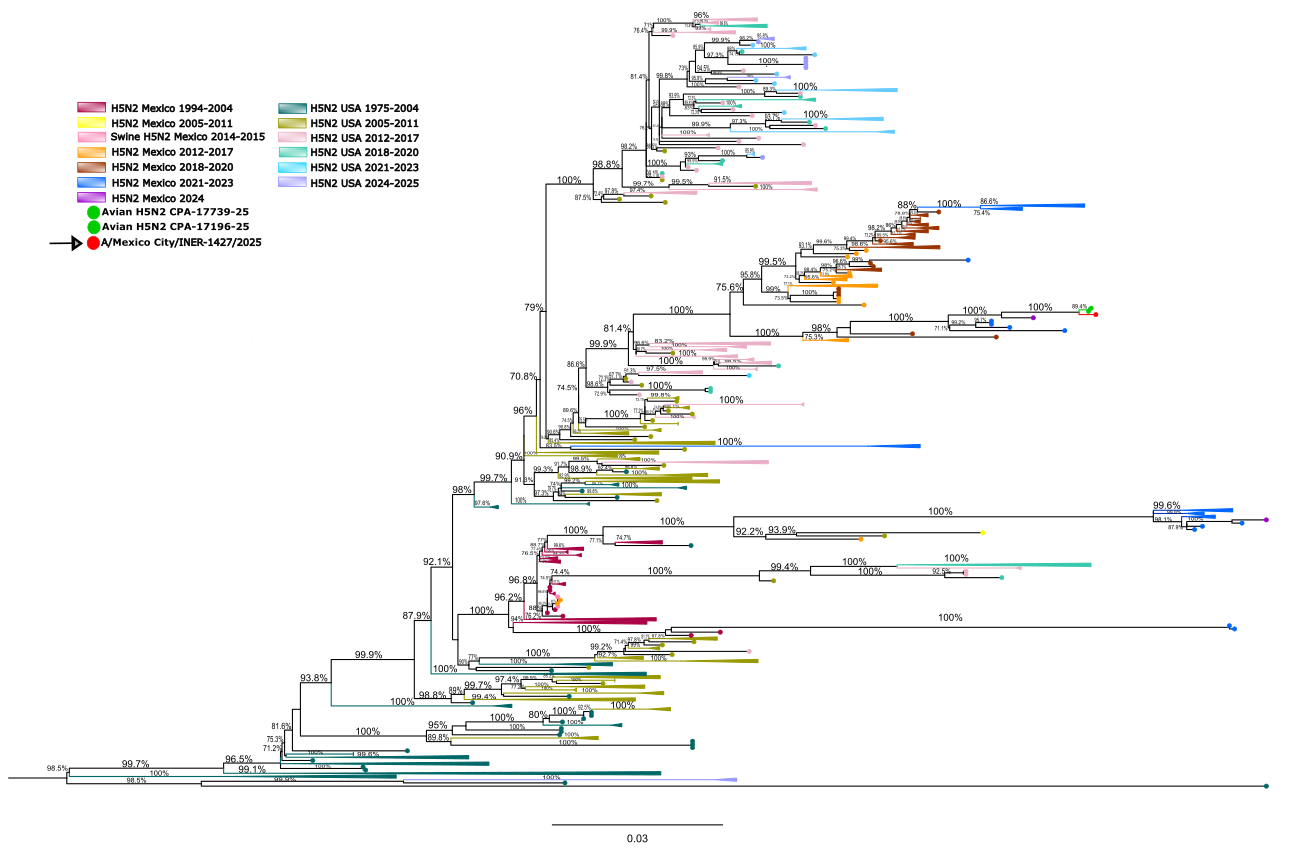

### Figure S3A and B. Maximum likelihood (ML) phylogenetic trees for PB1 (A) and NP (B) influenza H5N2 genetic segments. ML trees from 225 avian influenza

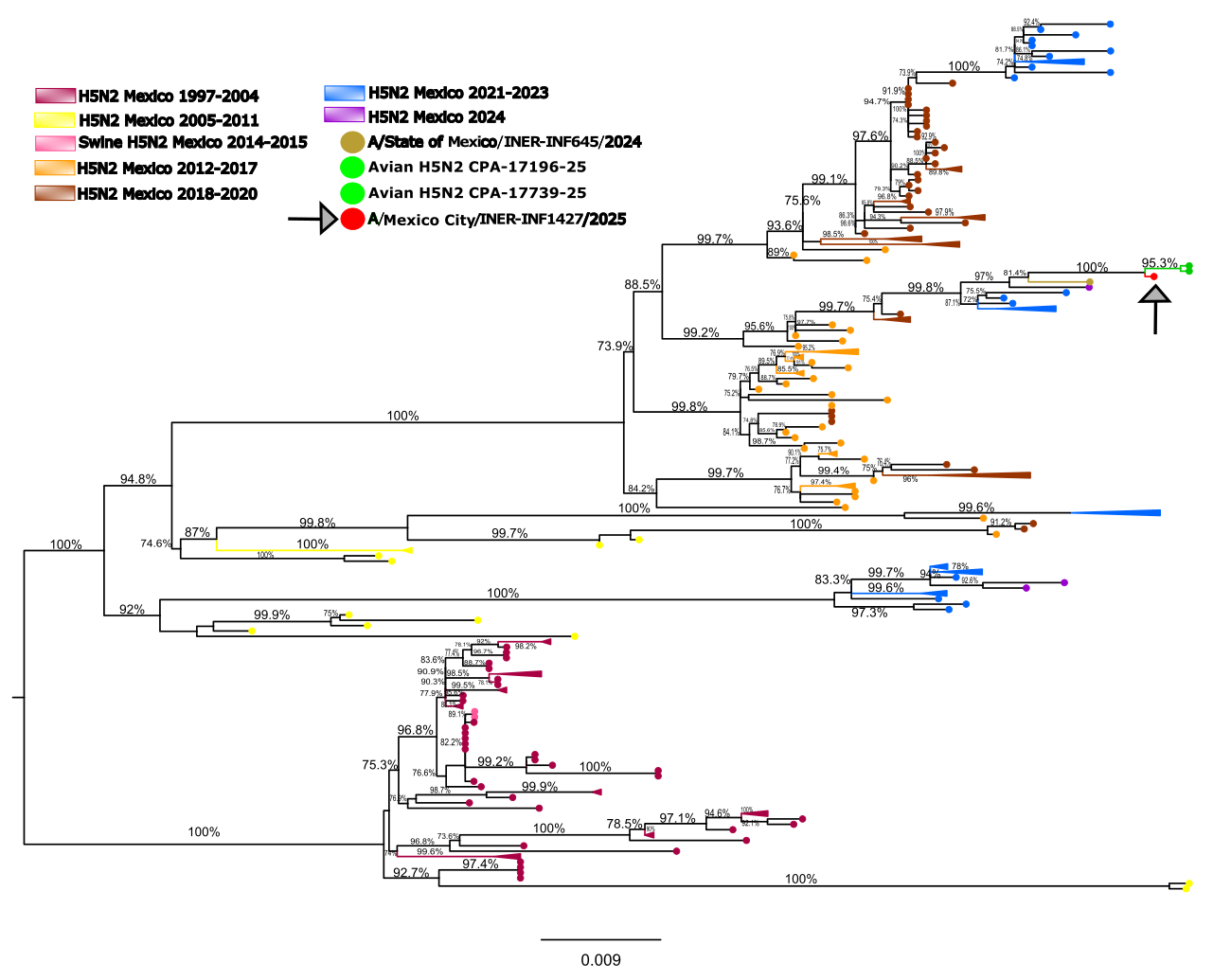
